# Subclinical variations on ECG and their associations with structural brain aging networks

**DOI:** 10.1101/2024.03.18.24304486

**Authors:** Elizabeth Haddad, William Matloff, Gilsoon Park, Mengting Liu, Neda Jahanshad, Ho Sung Kim

## Abstract

Impaired cardiac function is associated with cognitive impairment and brain imaging features of aging. Cardiac arrhythmias, including atrial fibrillation, are implicated in clinical and subclinical brain injuries. Even in the absence of a clinical diagnosis, subclinical or prodromal substrates of arrhythmias, including an abnormally long or short P-wave duration (PWD), a measure associated with atrial abnormalities, have been associated with stroke and cognitive decline. However, the extent to which PWD has subclinical influences on overall aging patterns of the brain is not clearly understood. Here, using neuroimaging and ECG data from the UK Biobank, we use a novel regional “brain age” method to identify the brain aging networks associated with abnormal PWD. We find that PWD is inversely associated with accelerated brain aging in the sensorimotor, frontoparietal, ventral attention, and dorsal attention networks, even in the absence of overt cardiac diseases. These findings suggest that detrimental aging outcomes may result from subclinically abnormal PWD.

## Introduction

The brain accounts for only about 2% of the body’s mass, yet it is estimated to receive 12% of cardiac output, and use 20% of the body’s oxygen (Williams and Leggett 1989, Raichle and Gusnard 2002). Given that this cardiac output provides the means by which cerebral blood flow is able to perfuse brain tissue, inevitably the heart and brain are intricately linked. A growing body of literature suggests that cardiovascular diseases (CVDs), including atrial fibrillation (AF), heart failure, and coronary artery disease, are associated with cognitive decline and brain imaging-derived features of aging (Tublin et al. 2019, Friedman et al. 2014).

P-wave indices on an electrocardiogram (ECG), specifically abnormalities in P-wave duration (PWD), offer valuable information of the electrical activity within the heart’s atria. The PWD represents the current moving from the sinoatrial node to the atrioventricular node and characterizes atrial depolarization (Lip et al. 2016). Anomalies in PWD, whether shorter or longer than the norm, are believed to reflect structural or physiological alterations in the left or right atria. These subtle variations thus provide markers of cardiac pathology and can serve as early indicators of various cardiovascular diseases, including AF and other arrhythmias, cardiovascular risk factors and related deaths (He et al. 2017, Nielsen et al. 2015, Magnani et al. 2009, Kosar et al. 2008, Uyarel et al. 2005, Magnani et al. 2015), and more recently dementia (Gutierrez et al., 2019) and cerebrovascular-related injuries (Reyes et al. 2023).

As ECG is more readily available and cheaper to obtain than MRI, charting a relationship between ECG outputs and brain health may help identify objective clinical risk factors for individuals at risk for faster brain aging even without a brain scan. Here, in a subset of UK Biobank participants who have both ECG and brain MRI available (Miller et al. 2016), we aim to identify if and how abnormal PWD may relate to accelerated brain aging, even in the absence of overt cardiac diseases. In particular, we use an advanced deep learning method capable of estimating “regional brain age” to characterize brain structural vulnerability related to atrial remodeling/dysfunction as measured by abnormal PWD.

## Results

### Demographic Characteristics

The 12,762 subjects selected for analysis were 52.6% female with a mean age and standard deviation of 63±7.36 years. The subset without major cardiac conditions (N=11,771) were 54.2% female with a mean age and standard deviation of 62.7±7.33. Demographic characteristics of all subjects stratified by percentiles of PWD may be found in **Table 1**. In this dataset, the median percentile (40-60th) corresponded to a PWD of 96-100ms. In general, subjects with low PWD were more likely to be female, whereas those with high PWD were more likely to be male (Wilcoxon test, *W*=16987621, *p* < 0.0001) (**Figure 1**). A higher incidence of AF, CAD, HF, and chronic kidney disease was observed in the lowest (<5%) and highest (>95%) percentiles.

**Figure 1.**
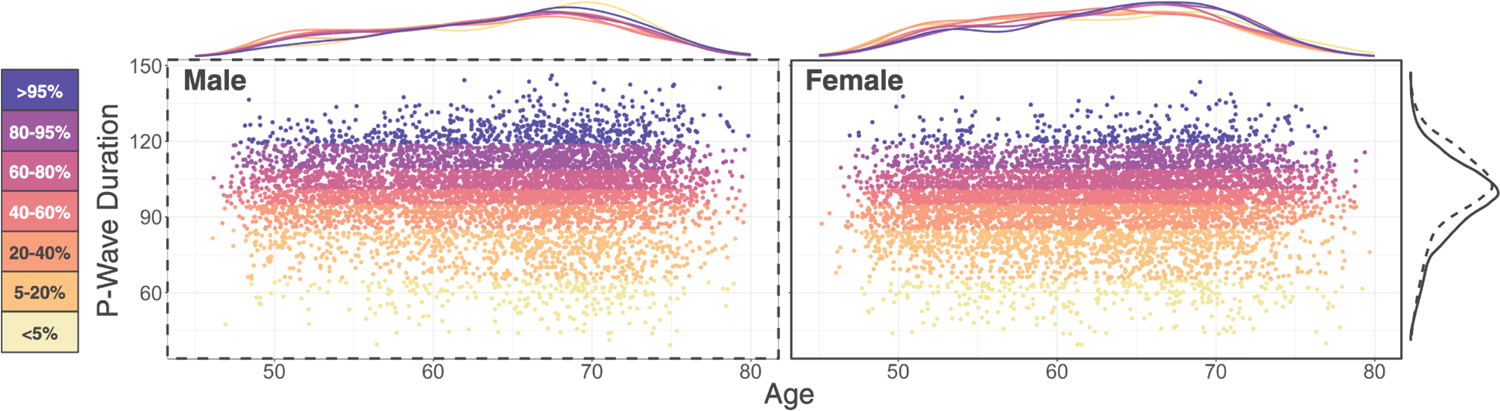
P-wave duration plotted against age and stratified by sex. Colors denote P-wave duration percentiles listed on the left hand side. Density plots across ages for each sex are depicted atop the scatters. Density plot on the right displays P-wave duration for males (dashed line) and females (solid line). Note higher densities of both high and low P-wave duration at older ages and across both sexes. Males tended to have longer P-wave duration whereas females favored shorter.

**Table 1.** Demographic characteristics stratified by percentiles of P-wave duration with associated interval limits listed.

| Characteristic | <5th<br>(40-64ms)<br>N = 560<sup>†</sup> | 5-20th<br>(66-84ms)<br>N = 1892<sup>†</sup> | 20-40th<br>(86-94ms)<br>N = 2325<sup>†</sup> | 40-60th<br>(96-100ms)<br>N = 2256<sup>†</sup> | 60-80th<br>(102-108ms)<br>N = 2732<sup>†</sup> | 80-95th<br>(110-118ms)<br>N = 2112<sup>†</sup> | >95th<br>(120-146ms)<br>N = 885<sup>†</sup> | Overall<br>N = 12762<sup>†</sup> |
| --- | --- | --- | --- | --- | --- | --- | --- | --- |
| Age | 63.95 [7.56] | 62.49 [7.46] | 62.13 [7.54] | 62.59 [7.24] | 63.15 [7.28] | 63.64 [7.20] | 64.42 [7.06] | 62.97 [7.36] |
| Female | 308 (55%) | 1,143 (60%) | 1,402 (60%) | 1,232 (55%) | 1,415 (52%) | 910 (43%) | 304 (34%) | 6,714 (53%) |
| Male | 252 (45%) | 749 (40%) | 923 (40%) | 1,024 (45%) | 1,317 (48%) | 1,202 (57%) | 581 (66%) | 6,048 (47%) |
| College | 232 (41%) | 783 (41%) | 983 (42%) | 1,009 (45%) | 1,220 (45%) | 983 (47%) | 422 (48%) | 5,632 (44%) |
| Coronary Artery Disease | 38 (6.8%) | 108 (5.7%) | 113 (4.9%) | 94 (4.2%) | 139 (5.1%) | 130 (6.2%) | 97 (11%) | 719 (5.6%) |
| Heart Failure | 4 (0.7%) | 8 (0.4%) | 14 (0.6%) | 12 (0.5%) | 10 (0.4%) | 15 (0.7%) | 11 (1.2%) | 74 (0.6%) |
| Chronic Kidney Disease | 7 (1.3%) | 14 (0.7%) | 11 (0.5%) | 12 (0.5%) | 19 (0.7%) | 16 (0.8%) | 14 (1.6%) | 93 (0.7%) |
| Atrial Fibrillation | 17 (3.0%) | 45 (2.4%) | 45 (1.9%) | 30 (1.3%) | 43 (1.6%) | 51 (2.4%) | 34 (3.8%) | 265 (2.1%) |
| Hypertension | 156 (28%) | 441 (23%) | 522 (22%) | 506 (22%) | 756 (28%) | 622 (29%) | 314 (35%) | 3,317 (26%) |
| Hypercholesterolemia | 84 (15%) | 280 (15%) | 337 (14%) | 335 (15%) | 428 (16%) | 386 (18%) | 208 (24%) | 2,058 (16%) |
| Diabetes | 33 (5.9%) | 95 (5.0%) | 108 (4.6%) | 99 (4.4%) | 156 (5.7%) | 119 (5.6%) | 64 (7.2%) | 674 (5.3%) |
| Sleep Apnea | 5 (0.9%) | 19 (1.0%) | 25 (1.1%) | 13 (0.6%) | 24 (0.9%) | 24 (1.1%) | 12 (1.4%) | 122 (1.0%) |
| On Hypertension Medication | 135 (24%) | 379 (20%) | 439 (19%) | 406 (18%) | 633 (23%) | 549 (26%) | 277 (31%) | 2,818 (22%) |
| On Cholesterol Medication | 131 (23%) | 380 (20%) | 444 (19%) | 428 (19%) | 575 (21%) | 513 (24%) | 269 (30%) | 2,740 (21%) |
| Systolic Blood Pressure | 136.12 [17.67] | 134.82 [17.2] | 135.62 [18.00] | 136.17 [17.46] | 137.59 [17.89] | 138.76 [17.72] | 139.11 [17.70] | 136.80 [17.74] |
| Diastolic Blood Pressure | 78.04 [9.45] | 77.79 [9.69] | 78.43 [9.95] | 78.35 [9.78] | 79.13 [10.22] | 79.41 [9.57] | 79.62 [10.27] | 78.70 [9.90] |
| Bone Mineral Density T-score | -0.23 [1.25] | -0.22 [1.20] | -0.33 [1.12] | -0.27 [1.13] | -0.24 [1.18] | -0.19 [1.19] | -0.04 [1.27] | -0.24 [1.18] |
| Body Mass Index | 26.89 [4.73] | 26.54 [4.62] | 26.26 [4.30] | 26.18 [4.21] | 26.59 [4.29] | 26.90 [4.27] | 27.50 [4.34] | 26.58 [4.36] |
| Smoking Status |  |  |  |  |  |  |  |  |
| Never | 347 (62%) | 1,212 (64%) | 1,495 (64%) | 1,460 (65%) | 1,692 (62%) | 1,304 (62%) | 529 (60%) | 8,039 (63%) |
| Previous | 192 (34%) | 601 (32%) | 711 (31%) | 715 (32%) | 916 (34%) | 732 (35%) | 314 (35%) | 4,181 (33%) |
| Current | 21 (3.8%) | 79 (4.2%) | 119 (5.1%) | 81 (3.6%) | 124 (4.5%) | 76 (3.6%) | 42 (4.7%) | 542 (4.2%) |
| Alcohol Status |  |  |  |  |  |  |  |  |
| Infrequent | 153 (27%) | 546 (29%) | 671 (29%) | 613 (27%) | 720 (26%) | 530 (25%) | 194 (22%) | 3,427 (27%) |
| Occasional | 321 (57%) | 1,037 (55%) | 1,283 (55%) | 1,258 (56%) | 1,512 (55%) | 1,167 (55%) | 518 (59%) | 7,096 (56%) |
| Frequent | 86 (15%) | 309 (16%) | 371 (16%) | 385 (17%) | 500 (18%) | 415 (20%) | 173 (20%) | 2,239 (18%) |
| ApoE4 copies |  |  |  |  |  |  |  |  |
| 0 | 418 (75%) | 1,372 (73%) | 1,673 (72%) | 1,640 (73%) | 1,975 (72%) | 1,516 (72%) | 652 (74%) | 9,246 (72%) |
| 1 | 130 (23%) | 477 (25%) | 601 (26%) | 563 (25%) | 690 (25%) | 545 (26%) | 220 (25%) | 3,226 (25%) |
| 2 | 12 (2.1%) | 43 (2.3%) | 51 (2.2%) | 53 (2.3%) | 67 (2.5%) | 51 (2.4%) | 13 (1.5%) | 290 (2.3%) |

<sup>†</sup>Mean [SD]; n (%)

### PWD and Regional Brain Aging

Regional BAI indices were negatively associated with PWD (i.e., shorter PWD associated with higher BAI). Indices that survived multiple comparisons testing included the sensorimotor (full set: *t* = −3.06, *q* = 0.007; CVD-control subset: *t* = −2.64, *q* = 0.025), fronto-parietal (full set: *t* = −3.53, *q* = 0.001; CVD-control subset: *t* = −3.07, *q* = 0.010), dorsal attention (full set: *t* = −4.11, *q* < 0.001; CVD-control subset: *t* = −3.67, *q* = 0.003), ventral attention (full set: *t* = −3.75, *q* = 0.001; CVD-control subset: *t* = −3.03, *q* = 0.010), and the language networks (full set: *t* = −2.92, *q* = 0.003; CVD-control subset not significant) (**Figures 2 & S3**).

**Figure 2.**
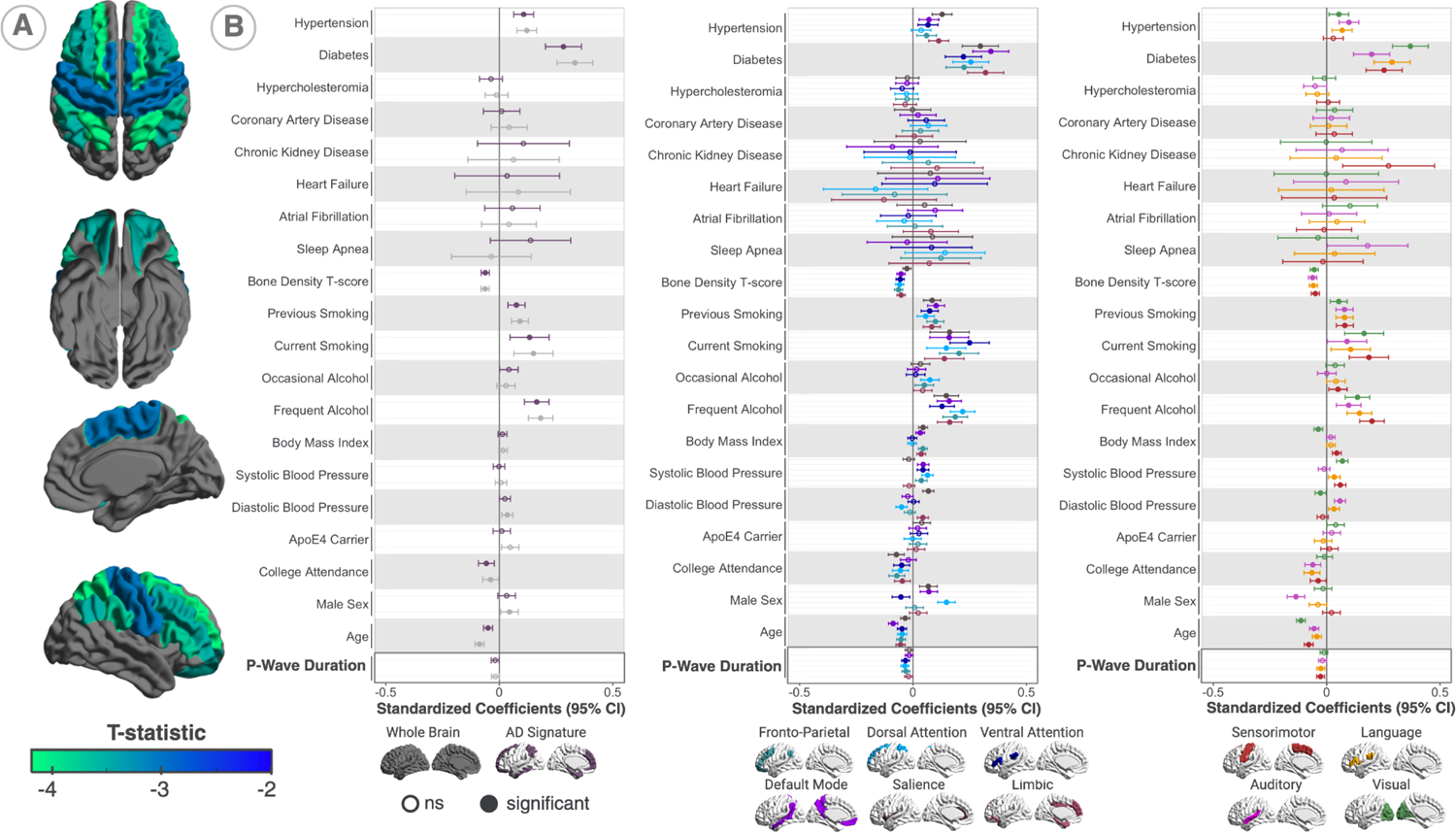
A. T-statistics of regional BAI (overlapping) associated with PWD. **B.** Standardized coefficients of PWD and all other regression coefficients (with the exception of the four MRI PCA components) from all 12 models (whole brain BAI, AD signature region BAI, and 10 regional BAI). Shorter P-wave duration was associated with a higher brain age. Regional BAI that survived multiple comparisons included the fronto-parietal, dorsal attention, and ventral attention (middle panel), as well as the sensorimotor and language networks (right panel). Covariates associated with a higher BAI include hypertension, diabetes, smoking status, and frequent alcohol consumption. Covariates associated with a lower BAI include a higher bone mineral density T-score, having a college education, and younger age.

The factor that showed the highest BAI for all 12 networks in all subjects and the CVD-control subset was diabetes. Hypertension and systolic blood pressure were also associated with a higher BAI for most regions. Other factors associated with a higher BAI for most, if not all, regional BAI included currently or previously smoking compared to never having smoked and frequent alcohol consumption compared to infrequent consumption. Factors associated with a younger than actual brain age included a higher bone mineral density T-score, having a college education, and lower chronological age. Biological sex showed some regional variability where compared to females, males tended to have a higher BAI in the salience, default mode, and dorsal attention networks. Conversely, males tended to have lower BAI in the ventral attention and auditory networks compared to females.

Models including a quadratic PWD did not show any significant associations. Models including PWD interactions with sex and atrial fibrillation similarly did not reveal any significant interactions.

### Regional Brain Aging and Cognition

The total number of subjects available for cognitive analysis from our initial set amounted to N=4,620 for average memory, N=4,447 for executive cognition, N=4,623 for processing speed, N=4,594 for reasoning, and N=4,401 for total cognition. All cognitive measures were associated with all regional BAI for executive function, processing speed, reasoning, and total cognition with the exception of the ventral attention network for executive function and the dorsal attention network for reasoning (**Figure 3**). Average memory showed less robust associations, but was still significantly associated with whole brain, default mode, salience, and visual BAI.

**Figure 3.**
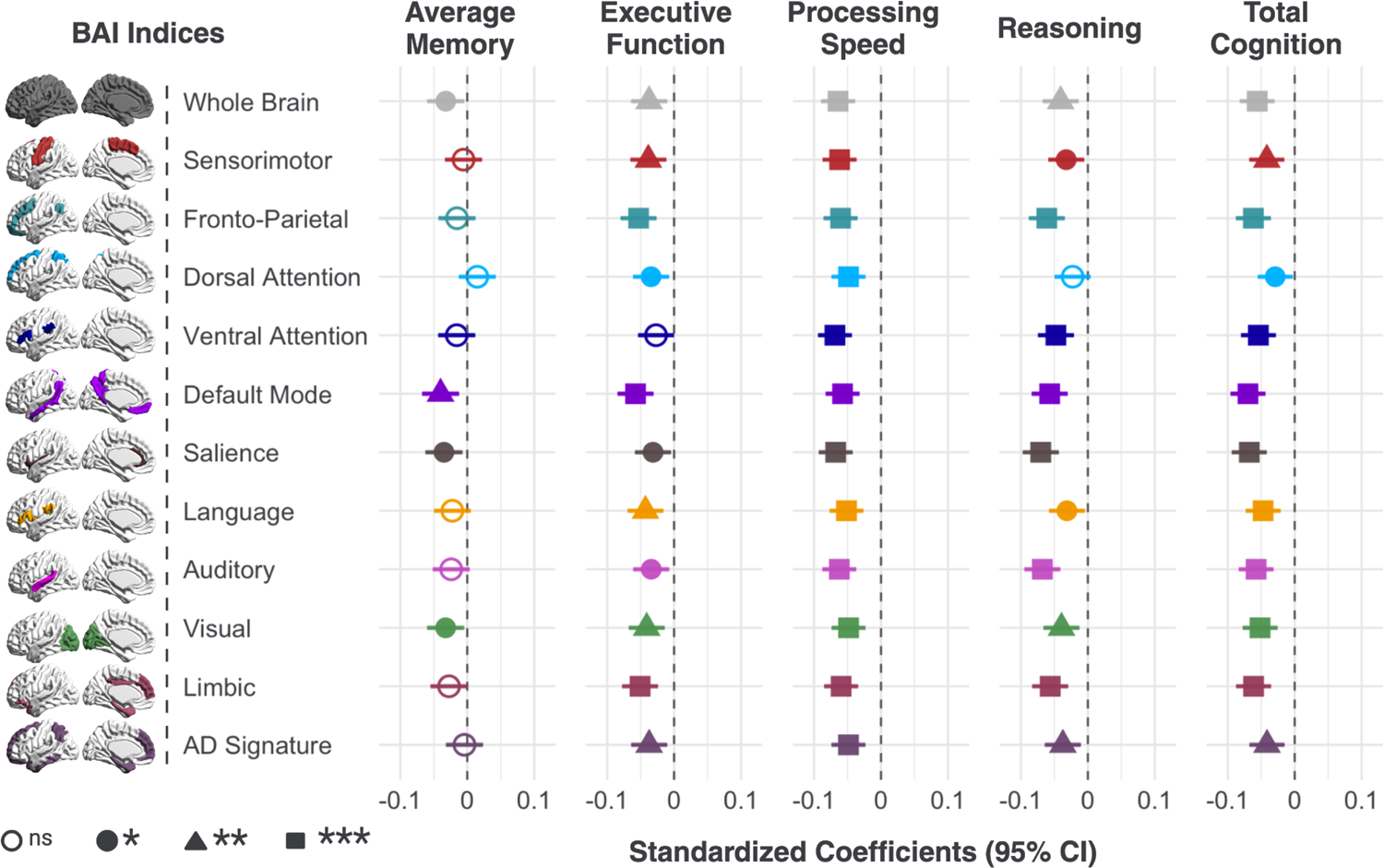
Regional BAI associations with composite cognitive measures – average memory, executive function, processing speed, reasoning, and total cognition. Values displayed are standardized regression coefficients. All models included age, sex, and college attendance as covariates. Higher cognitive scores were generally associated with a lower brain age.

### PWD and Cognition

None of the composite cognitive measures were associated with PWD after correcting for age, sex, and college education.

## Discussion

Our study aimed to characterize the brain aging patterns and cognitive outcomes associated with PWD, an ECG-surrogate of underlying atrial function. We found the following:

1. A negative association between PWD and distinct patterns of regional brain aging exists. Several brain networks, including the sensorimotor, frontoparietal, ventral attention, dorsal attention, and language networks, were particularly vulnerable to short PWD. These networks were associated with global cognition and also specific cognitive measures including executive function, processing speed, and reasoning.
2. Our findings exist even when adjusting for cardiovascular risk factors, and when excluding major cardiac conditions (with the exception of the language network), suggesting detrimental aging outcomes may result from subclinically abnormal PWD.
3. We did not find direct associations between PWD and cognition, but did find associations between cognition and brain age measures. This suggests abnormal PWD may exert its effects on the brain in advance of detectable cognitive decline, but longitudinal studies are needed to confirm this hypothesis.

Abnormal PWD is thought to reflect underlying atrial abnormalities, which can include atrial dilation, atrial muscular hypertrophy, elevated atrial pressure, impaired ventricular distensibility, or delayed intra/inter atrial contraction (Hancock et al. 2009). Prolonged PWD, typically defined as greater than 120ms, is often a result of inter-atrial block (Power et al., 2022). Although less is known about the correlates of abnormally short PWD, in certain instances, short PWD may be considered an earlier and more subtle feature than prolonged PWD. It has been hypothesized that shortened atrial repolarization and refractory periods could indicate changes in atrial electrical properties preceding more overt abnormalities associated with prolonged PWD, which can eventually result in arrhythmogenesis (Nielsen et al. 2015, Zhou et al. 2023). Recent research has started to underscore that shorter PWD is also associated with adverse outcomes, including AF, heart failure, stroke, and dementia (Ostrowska et al. 2022, Ostrowska et al. 2022, Chen et al. 2022, Zhou et al. 2023).

Disruptions in atrial function could theoretically impact cerebrovascular health through hemodynamic dysregulation, cerebral hypoperfusion, systemic inflammation, and hypercoagulation (van der Velpen et al. 2017), but research related to the specific contribution of PWD is scant. It is clear, however, that gradual disruptions of cerebrovascular hemodynamics can be a major contributor to the initiation of vascular dementia pathology. Subclinical decreases in cardiac function and increases in arterial stiffness can lead to cerebral hypoperfusion, blood brain barrier damage, and detrimental pulsatile blood flow. The resulting neurovascular dysfunction and inflammation can impair glymphatic clearance and exacerbate existing AD pathology leading to synaptic dysfunction and eventually brain atrophy (Moore and Jefferson 2021). A recent study by (Reyes et al. 2023) found an association between prolonged PWD and various cerebrovascular related injuries, but not with gross lobar volumes. We, on the other hand, found a significant association between short PWD and accelerated brain aging. Our enhanced sensitivity may be explained by the fact that we used a specific component of volume (i.e. thickness) as well as grey/white matter intensity ratio with a finer parcellation of brain networks as opposed to volume measurement only in the whole brain and cortical lobes. While our results also suggest that longer PWD is associated with lower brain age given the lack of significance in the models we tested with a quadratic PWD term, this may be a result of fewer participants in the UK Biobank having what is often defined as “prolonged PWD”.

We did not *a priori* define abnormal PWD as dichotomous – prolonged or not – and instead investigated these associations in an unbiased manner using the continuous measure, enabling the analysis of bidirectional PWD abnormalities (short and prolonged) without a threshold. Some studies prefer to use percentile cutoffs to circumvent assumptions, although admittedly these can vary widely from population to population and by other factors including age, sex, and ethnicity (Soliman et al. 2013, Nielsen et al. 2015). For example, Nielsen et al. (2015) assessed a large primary care population from the Copenhagen ECG Study and established that the optimal PWD with respect to the lowest risk of AF was 100-105 ms which fell between the 49th and 61th percentile in our study. Abnormally short PWD in their study, or <5th percentile, was less than 90 ms compared to less than 65 ms in our study. Prolonged PWD in their study, or >95th percentile, was greater than 131 ms compared to greater than 119 ms in our study. Given the differences in percentile cutoffs across populations, we opted to use a continuous PWD. Nonetheless, Nielsen et al. found an elevated risk of AF (and cardiovascular death and stroke although not as robust) in those with abnormally short *and* long PWD. Using a “stopped” Cox model, they also showed that the association between shorter PWD and AF risk was slightly stronger when evaluating short term effects compared to long term. They hypothesized that short PWD, resulting in a more rapid condition time, may be a substrate for the early development of AF. While it is possible that the participants with abnormally short PWD in our study will go on to develop AF, a longitudinal study design is needed to evaluate this.

Although we detected associations between brain structure and PWD & brain structure and cognition, we did not observe a direct association between PWD and cognition. This parallels previous longitudinal work from the ARIC-NCS study which showed that abnormal PWD increased the risk of dementia, but did not reduce cognitive scores. The authors suggest that atrial abnormalities may increase the risk of sudden, but major, events such as stroke, as opposed to a more subtle and progressive decline (Gutierrez et al. 2019). While this may be true, this study did not investigate the effects of abnormally short PWD, and instead only investigated prolonged PWD associations with cognition. The brain areas that we found to be associated with PWD include the frontoparietal network responsible for executive control, the sensorimotor network in charge of processing bodily sensations and executing appropriate motor responses, and the attentional networks (dorsal and ventral) responsible for top down and bottom up processing respectively (Petersen and Posner 2012). These networks are all in communication with one another to facilitate various aspects of cognitive function tested in our study. Our results may suggest that subtle brain alterations due to short PWD may impose accelerated aging effects to structural networks that are responsible for important cognition tasks, but that these occur in advance of detectable cognitive decline.

We note, however, that our study is cross-sectional in nature and a longitudinal study design is needed to investigate whether short PWD predicts future cognitive deficits. While we attempted to account for all possible confounders by including them as covariates in our linear models, we note that there may be other contributing variables that we could not account for due to our retrospective study design. Another limitation we acknowledge is that our model only includes cortical features, even though subcortical/deep gray matter and white matter features may be important predictors of brain aging, particularly those vulnerable to vascular pathology (Wardlaw et al. 2013). Future work will continue to investigate these associations. Lastly, it may also be the case that the relationship between short PWD and cognitive decline is relatively weak in the preclinical population we are studying, and that effects may be more pronounced in a clinical population of AD and related dementia cases.

Nonetheless, we find, even in the absence of major cardiac conditions and controlling for cardiovascular risk factors, that PWD still remains a significant predictor of many regional BAI. This suggests that early detection and treatment of poor cardiac function, prior to manifestation of arrhythmias, may help curb accelerated brain aging. Moreover, these associations may be helpful in understanding how the heart-brain axis impacts brain health and cognitive aging trajectories.

## Online Methods

### Study Population

The UK Biobank is a large population-based study that has collected deep phenotypic and genetic data from approximately 500,000 community dwelling adults in the UK (Bycroft et al. 2018). Its dense phenotyping has resulted in extensive information on health status and lifestyle, in addition to the collection of biological, physical, and cognitive assessments. Here, data from 40,678 participants with imaging and ECG were initially considered for this study. 6,778 ECG diagnoses related to poor readings were excluded, resulting in a total N=33,900. Subjects with neurological disorders, putative sex chromosome aneuploidy, excess relatives, and missing covariates were also excluded (N=29,364). A total of 12,819 of these subjects had our outcome variable, regional brain age, available. Lastly, to assess subclinical effects of PWD on brain aging, subjects with ECG readings indicating an acute myocardial infarction or bifascicular block were excluded. This resulted in a total of 12,762 subjects for analysis. To assess if abnormal PWD affects regional brain structure even in the absence of overt cardiac-related diseases, we performed the same analysis in a CVD-control subset for which we excluded subjects with AF, CAD, HF, and chronic kidney disease – resulting in a subset total of 11,771. Conditions were categorized as in Khurshid et al. (2018). Details about inclusion/exclusion criteria are provided in **Figure S1**.

### MRI Acquisition and Preprocessing

We used the CIVET pipeline (Ad-Dabbagh et al., 2006) on T1-weighted brain MRI to extract cortical features including thickness and gray/white matter intensity. This pipeline includes the following serial steps: non-uniform intensity correction (Sled et al. 1998), brain extraction (Smith 2002), registration to a stereotaxic space (Collins et al. 1994), brain tissue segmentation (Zijdenbos et al. 1998), and reconstruction of inner and outer cortical surfaces (Kim et al. 2005), resulting in 40,962 vertices in each hemisphere. These cortical surface models underwent an iterative surface registration process to ensure optimal correspondence at each vertex across individuals (MacDonald et al., 2000), (Lyttelton et al., 2007). Cortical thickness measurements were obtained by calculating the Euclidean distance between the vertices of the inner cortical surface and their corresponding vertices on the outer cortical surface. The GM/WM intensity ratio information was derived from the inner surface (Lewis et al., 2018).

### Regional Brain Age Index (BAI)

To develop predictive models for regional brain age indices (BAI), we divided the cortical surface into ten functional subregions based on Yeo et al. (2011) and the AAL cortical parcellation atlas (Tzourio-Mazoyer et al., 2002). These subregions include the sensorimotor, frontoparietal, dorsal attention, ventral attention, default mode, salience, language, auditory, visual, and limbic networks. Two additional regions have been defined – one based on the total cortical thickness and the other based on the cortical regions associated with Alzheimer’s disease (AD signature region) (Dickerson et al., 2009). Regional brain ages were extracted using Graph Convolutional Networks (GCNs) (Defferrard et al., 2016, Shuman et al., 2013), exploiting the graph structure of the data. Cortical features such as cortical thickness (Thambisetty et al., 2010) and GM/WM intensity ratio (Putcha et al., 2023) served as the signal at each node. GCNs used graph Fourier transforms, filtering, and pooling operations for feature aggregation. The GCN architecture included a graph convolutional layer, a ReLU activation function, a graph max pooling operation, and a fully connected layer for brain age prediction. The overall flow of the GCNs model is shown in **Figure S2**. The training process involved mean square error as the loss function, the Adam optimizer, 800 epochs, a learning rate of 10e-6, L2 regularization to prevent overfitting, and a batch size of 2. A 5-fold cross-validation was performed using 17,791 individuals of European ancestry (52.7% female with a mean age and standard deviation of 63.15±7.42 years) who were considered to be neurologically healthy as defined by ICD and self report, resulting in an ensemble of five trained models. BAIs were calculated by subtracting the chronological age from the predicted brain age. A positive BAI indicates accelerated aging, while a negative BAI suggests decelerated aging. To address regression dilution bias, we used the linear trend removal method proposed by Smith et al., (2019). Linear trend removal involves regressing the trend of BAIs on age to obtain corrected BAIs, eliminating age-related bias and ensuring relative brain health status independent of age.

### Cognitive Outcomes

Composite averages were calculated for four cognitive domains: memory, executive function, processing speed, and reasoning. Memory was assessed using numeric memory and paired associate learning tasks. Executive function was assessed using the trail making A and B tasks in addition to tower rearranging. Processing speed used reaction times and symbol digit substitution and lastly, reasoning used fluid intelligence and matrix pattern completion tasks. Respective data fields for each of these domains may be found in **Table S1**. All scores were standardized such that higher values indicate a better cognitive score (ex: reaction time was multiplied by −1). Values from all four of these categories were additionally averaged to test a fifth cognitive variable – total cognition.

### ECG Measures

Resting 12-lead ECGs and interval measurements were assessed using the Cardiosoft v6 program from GE Healthcare. The PWD was measured as the duration from P-Onset to P-Offset (https://biobank.ndph.ox.ac.uk/showcase/ukb/docs/CardiosoftFormatECG.pdf).

### Statistical Analysis

Principal component analysis (PCA) was run on mixed data, a combination of numerical and categorical variables related to scanner biases affecting the T1w image (Alfaro-Almagro et al., 2021), using the *PCAmixdata* package (Chavent et al., 2014). Linear regressions for each regional BAI were run with the following covariates: 4 MRI PCA components that cumulatively explained at least 80% of the variance, age, sex, college attendance, the presence of hypertension, diabetes, hypercholesterolemia, sleep apnea, heel bone mineral density T-score, smoking status, alcohol consumption, body mass index (BMI), ApoE4 carrier status, systolic blood pressure, diastolic blood pressure, and PWD. We chose to model continuous PWD so as to not bias our outcomes with previously defined “normal” ranges, as these largely vary across populations and by age, sex, and ethnicity (Soliman et al., 2013), (Nielsen et al., 2015). CAD, HF, AF, and chronic kidney disease were also included in the models with all subjects. Respective datafield IDs may be found in **Table S1**. Cognitive associations were performed between PWD and regional BAI. All models included age, sex, and college education as a binary variable. Benjamini & Hochberg correction for multiple testing was performed across all 10 regional BAI measures, whole brain, and AD signature BAI for the following: 1) regional BAI associations with PWD, 2) regional BAI associations with cognition, and 3) PWD associations with cognition. Exploratory analyses were also performed to test the inclusion of a quadratic PWD term as well as PWD interactions with AF and sex.

## Data Availability

UK Biobank data available here: https://www.ukbiobank.ac.uk/

## Supplemental Figures/Tables

**Figure S1.**
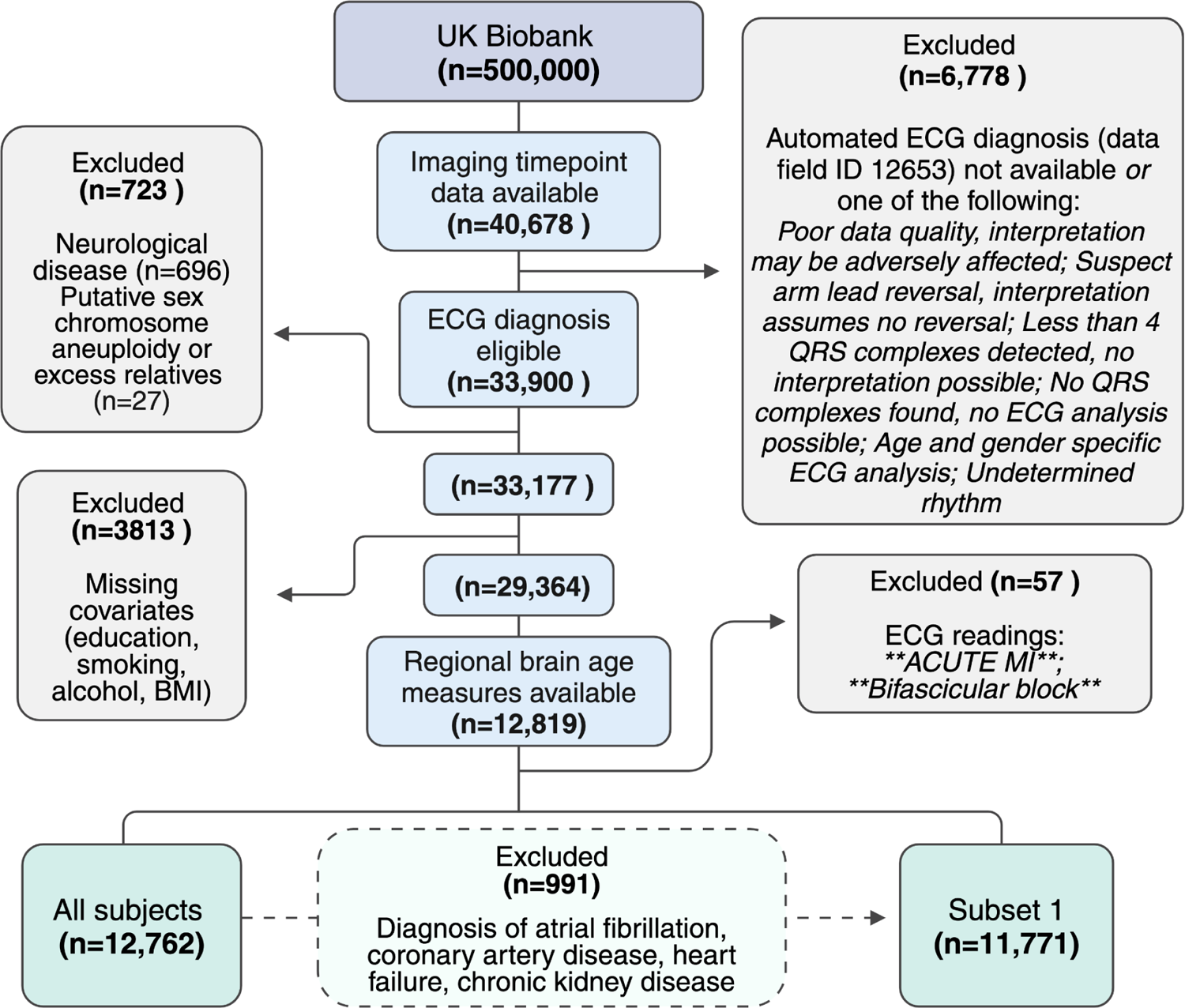
Inclusion flowchart. Self reported non-cancer illness codes (1082, 1083, 1240, 1244, 1245, 1246, 1247, 1256, 1258, 1259, 1261, 1262, 1263, 1264, 1266, 1289, 1397, 1425, 1433, 1659) and ICD10 codes for dementia and Parkinson disease.

**Figure S2.**
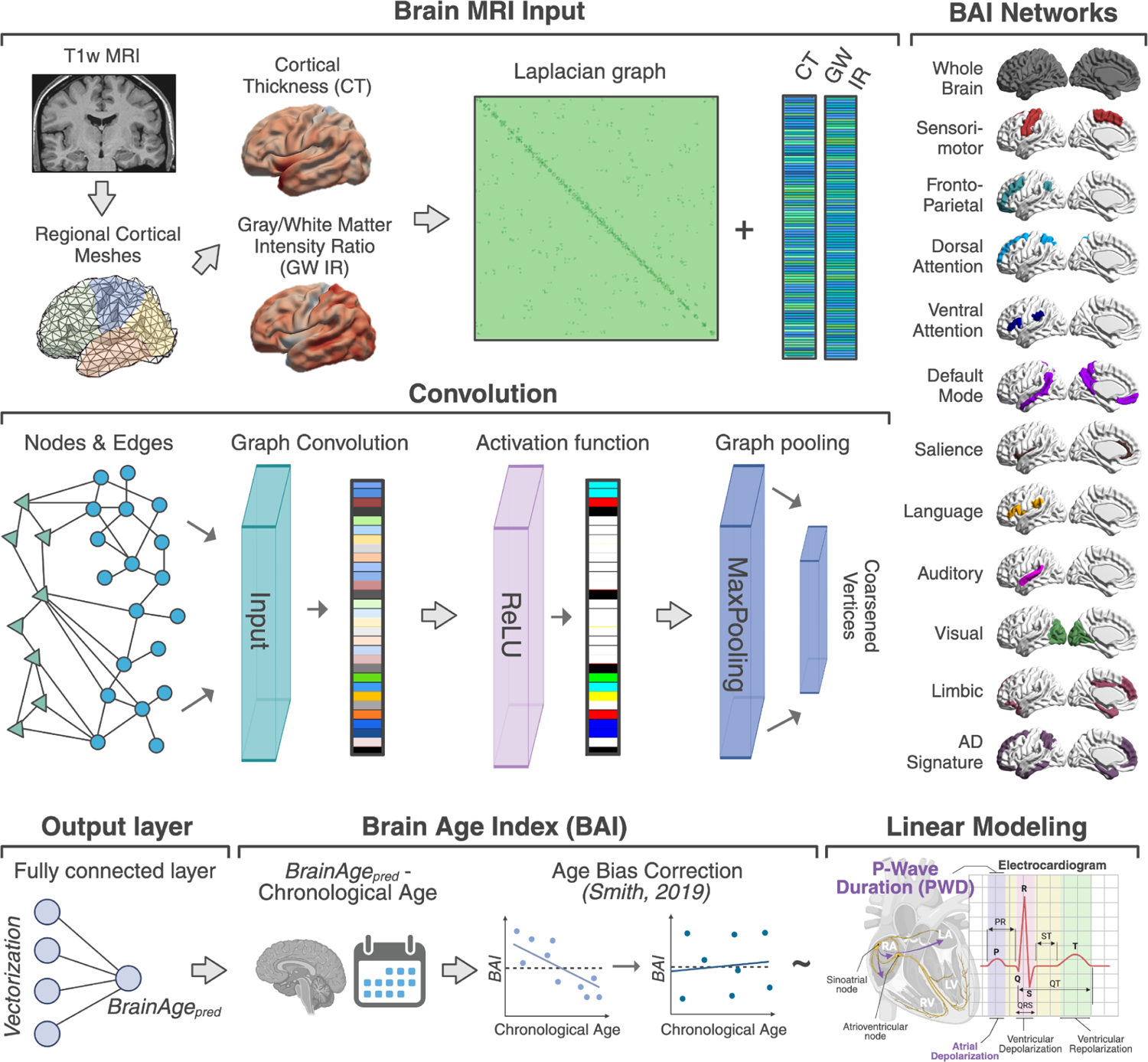
Methodological overview of BAI extraction. T1w brain aging features including cortical thickness and grey/white matter intensity ratio were inputs into a graph convolutional network resulting in a fully connected layer and predicted “brain age” outputs. Chronological age was subtracted from predicted brain age resulting in 12 regional brain age indices (BAI). Chronological age was regressed out from each BAI and used for analysis. Linear modeling was performed with PWD as a predictor variable while correcting for possible confounders by including them as covariates. PWD represents the current moving from the sinoatrial node to the atrioventricular node and characterizes atrial depolarization.

**Table S1.** Variables and respective data fields used for cardiovascular risk related covariates, cognition variables, and scanner/bias (MRI) used for principal component analysis. Dx: diagnosis; Tx: treatment. *Note:* Table Position was initially explored but due to high correlation with Brain Z Position (r_(12,762)_=-0.92, p <0.001), it was not included in the MRI PCA.

| **Cardiovascular Risk Variables** | **Datafield ID** |
| --- | --- |
| Hypertension Dx | 41270, 20002, 6150 |
| Hypercholesterolemia Dx | 41270, 20002 |
| Diabetes Dx/Tx | 41270, 20002, 6153, 6177 |
| Coronary Artery Disease Dx | 41270, 41272, 20002, 20004, 6150 |
| Heart Failure Dx | 41270, 20002 |
| Atrial Fibrillation Dx | 41270, 41272, 20002, 20004, 12653 |
| Chronic Kidney Disease Dx | 41270, 41272, 20002, 20004 |
| Sleep Apnea Dx | 41270, 20002 |
| Alcohol Consumption (infrequent, frequent, occasional) | 1558, 41270 |
| Smoking Status (never, previous, current) | 20116 |
| BMI | 23104 |
| Heel bone mineral density T-score | 4106, 4125 |
| APOE4 carrier | - |
| Systolic Blood Pressure | 4080, 93 |
| Diastolic Blood Pressure | 4079, 94 |
| **Cognitive Variables** | - |
| Memory | 4282, 20197 |
| Executive Function | 6348, 6350, 21004 |
| Processing Speed | 20023, 23324 |
| Reasoning | 20016, 6373 |
| **Scanner/Bias (MRI) Variables** | - |
| Scan Site | 54 |
| Brain X Position | 25756 |
| Brain Y Position | 25757 |
| Brain Z Position | 25758 |
| Inverse SNR | 25734 |
| Inverse CNR | 25735 |
| Linear Registration Discrepancy | 25731 |

**Figure S3.**
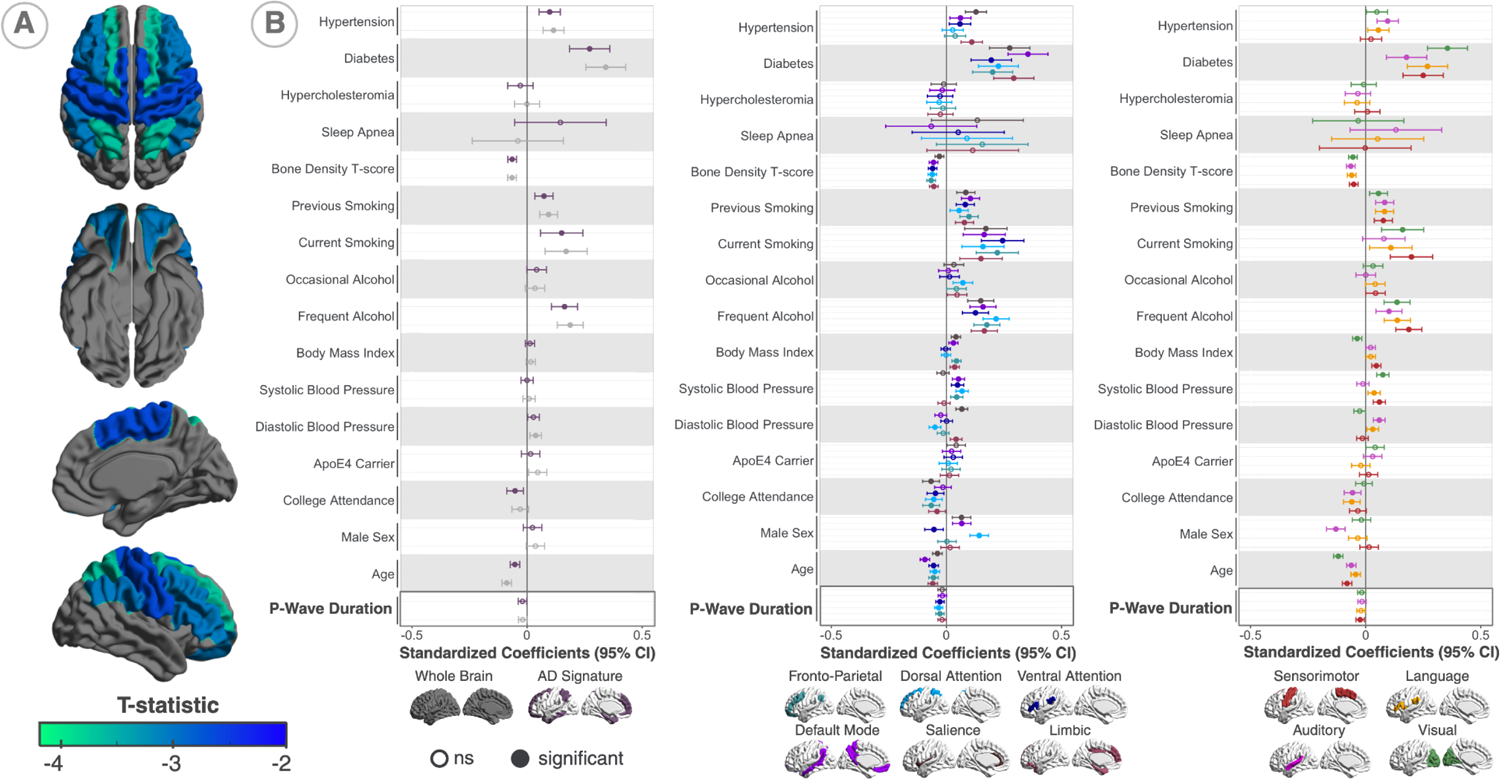
**A.** T-statistics of regional BAI (overlapping) associated with PWD in those without major cardiac conditions. **B.** Standardized coefficients of PWD and all other regression coefficients (with the exception of the four MRI PCA components) from all 12 models (whole brain BAI, AD signature region BAI, and 10 regional BAI). Shorter P-wave duration was associated with a higher brain age. Regional BAI that survived multiple comparisons included the fronto-parietal, dorsal attention, and ventral attention (middle panel), as well as the sensorimotor network (right panel). Covariates associated with a higher BAI include hypertension, diabetes, smoking status, and frequent alcohol consumption. Covariates associated with a lower BAI include a higher bone mineral density T-score, having a college education, and younger age.

## Acknowledgements

This work was supported in part by National Institutes of Health (R01AG059874), National Institutes of Health (P41EB015922), and Bright Focus Research Grant award (A2019052S). UK Biobank Resource under Application Number: 11559

